# Structural injuries are associated with better emotional wellbeing after traumatic brain injury

**DOI:** 10.1101/2025.07.24.25332156

**Authors:** Samuel B. Snider, Hui Shi, Chiara Maffei, Natalie Gilmore, Holly J. Freeman, Alexander S. Atalay, Jian Li, Jessica A. Yeager, Raj G. Kumar, Belinda Yew, Erin M. Conley, Ariana L. Velazquez, Julia Kirschenbaum, Enna Selmanovic, Holly Carrington, Yelena G. Bodien, Jeanne M. Hoffman, Christine Mac Donald, Brian L. Edlow, Kristen Dams-O’Connor

## Abstract

**Background:** Emotional disturbances are common after traumatic brain injury (TBI), yet the neuroanatomic basis of these sequalae remains unclear. We aimed to determine whether emotional wellbeing is associated with the extent and/or location of structural brain injury in individuals with chronic TBI.

**Methods:** We studied 188 adults with chronic (≥1 year post-injury) TBI enrolled in the multi-center Late Effects of TBI (LETBI) study. Participants underwent T1-weighted MRI to assess cortical lesion location and volume, and diffusion MRI to measure global mean white matter fractional anisotropy (FA), defined as the mean FA across 40 white matter tracts. The primary outcome was emotional wellbeing, measured as the RAND-36 emotional wellbeing subscale t score. To identify associations between structural injury and the primary outcome, we used multivariate linear models adjusted for age, sex, years of education, time since injury, history of psychiatric illness, and multiple indices of scan quality.

**Results:** The cohort was predominantly male (68%) and white (88%), with a mean age of 58 years (SD 15) and median time since injury of 8 years (IQR 14). The presence of cortical lesions was independently associated with greater emotional wellbeing scores (RAND-36 adjusted t score was 4.1; 95% CI: [1.0, 7.0] points higher in participants with versus without lesions; p=0.009). Right temporal and lateral orbitofrontal lesions associated with lower (worse) emotional wellbeing scores, and left hemispheric lesions associated with higher (better) scores. Lower global white matter FA was independently associated with greater emotional wellbeing scores (RAND-36 adjusted t score was 2.7 [0.8, 4.7] points higher per 1 SD decrease in FA; p = 0.008). The strongest associations between individual white matter tract integrity and emotional wellbeing were in the anterior (genu) corpus callosum (β = -3.0, [-1.3, -4.7]; pHolm=0.034) and the left cingulum bundle (β = -3.0, [-1.2, -4.8]; pHolm=0.048).

**Conclusions:** In a large cohort of participants with chronic TBI, structural brain injuries were associated with better emotional wellbeing scores. These findings challenge traditional interpretations of emotional disturbances after TBI and motivate future studies to determine whether specific structural brain injury locations may be causally associated with improvements in emotional wellbeing.

## Introduction

Emotional disturbances are a prevalent and frequently disabling sequelae of traumatic brain injury (TBI)^1–4^. Such disturbances—including depression, anxiety, irritability, and affective lability— can impede functional recovery and reduce quality of life^5^. Despite their high prevalence (20–50% for depression^6–8^) and clinical significance after TBI, it is unclear whether or how emotional disturbances are related to injury to specific brain structures or distributed networks. Studies seeking to measure the association between injury severity and specific mood or neurobehavioral symptoms have either produced null^7, 9, 10^ or conflicting^4, 5, 11–13^ results.

After a TBI, structural brain MRI typically reveals two primary types of pathology: encephalomalacic lesions, measurable on a standard T1-weighted sequence and representing the chronic sequelae of cortical contusions^14, 15^, and disrupted white matter integrity, quantified using diffusion MRI (dMRI)^16^ and representing the sequalae of diffuse axonal injury^17–19^. There have been limited a number of studies on the location of structural pathology after TBI and outcomes related to emotional wellbeing. In studies specifically focused on depression in chronic TBI, both cortical atrophy and reduced white matter integrity after TBI have been reported to be associated with depressive symptoms, but the implicated cortical regions and white matter tracts are inconsistent across studies^20^. Clarifying this relationship could help identify novel targets for neuromodulatory therapies^21–23^ in patients recovering from TBI.

A relatively new technique known as lesion network mapping^24^ has identified distributed networks of brain regions associated with depression^25, 26^, anxiety^27^, and post-traumatic stress disorder^28^—individual elements of emotional wellbeing—when affected by focal brain lesions. However, these networks were derived from patients with stroke or penetrating head trauma and their relevance to individuals with non-penetrating TBI, in whom the pathophysiological mechanisms of emotional disturbance may be distinct^29^, is uncertain.

Cognitive sequellae of structural injury (e.g., impaired attention, anosognosia), as well as factors not directly related to structural brain injury, may also influence emotional wellbeing after TBI. One of the strongest published predictors of emotional disturbances after TBI is a pre-morbid psychiatric history^5, 8^. Consequently, it remains unclear whether emotional disturbances after TBI predominantly arise from structural brain injury, premorbid psychological health, or complex interactions between both factors^30^.

In the current study, we investigated the neuroanatomic correlates of emotional wellbeing in individuals with chronic TBI from the Late Effects of TBI (LETBI) study—a comprehensive, multimodal investigation involving individuals evaluated at least one year post-injury.^31^ We focused on two structural MRI markers: encephalomalacic cortical lesions detected by T1-weighted MRI and white matter integrity measured using fractional anisotropy (FA) from dMRI. For each type of pathology we sought to: (1) determine whether the presence or extent of injury was associated with emotional wellbeing; and (2) use data-driven analyses to identify the neuroanatomic regions associated with emotional wellbeing.

## Methods

### Participants and Study Design

We performed a cross-sectional analysis of participants from the LETBI study^31^. LETBI is a multi-center, observational cohort that enrolls adults with chronic TBI (≥1 year post-injury) to undergo brain imaging and comprehensive neurobehavioral characterization. Detailed study inclusion and exclusion criteria have been previously published.^14, 31^ Study inclusion required self-report or medical record evidence of (a) head trauma resulting in Glasgow Coma Scale (GCS) score < 13 on emergency department arrival, loss of consciousness lasting at least 30 min, post-traumatic amnesia lasting over 24 h, or acute trauma-related intracranial neuroimaging abnormality. Exclusion criteria for LETBI included active neuropsychiatric conditions that could preclude study participation and/or confound assessments (e.g., active psychosis).

Among 301 patients enrolled across three United States academic medical centers between 2014 and 2022, the analytic cohort included participants who underwent study MRI and completed a neurobehavioral evaluation (N= 188; Table 1).

**Table 1:** Cohort Characteristics.

| Table 1: Cohort Characteristics |  |  |  |
| --- | --- | --- | --- |
|  | MRI and Outcomes (N=188) | Missing MRI or Outcomes (N=113) | Difference P Value |
| Mean Age (SD) | 58 (15) | 57 (18) | 0.57 |
| Male Sex (%) | 129/188 (69%) | 60/110* (55%) | 0.021 |
| White Race (%) | 166/188 (88%) | 92/110 (84%) | 0.46 |
| Attended College (%) | 128/188 (68%) | 86/108 (80%) | 0.045 |
| Marital Status (%)<br>*Never married*<br>*Married/partnered*<br>*Divorced/widowed* | 45/188 (24%)<br>83/188 (44%)<br>60/188 (32%) | 30/108 (28%)<br>47/108 (44%)<br>31/108 (29%) | 0.73 |
| Employment (%)<br>*Working/Student*<br>*Unemployed*<br>*Retired*<br>*Disabled*<br>*Other* | 63/188 (34%)<br>10/188 (5%)<br>56/188 (30%)<br>45/188 (24%)<br>14/188 (7%) | 33/108 (31%)<br>8/108 (7%)<br>32/108 (30%)<br>29/108 (27%)<br>6/108 (6%) | 0.87 |
| Median years since most recent TBI (IQR) | 8 (14) | 8 (15) | 0.50 |
| Median Glasgow Outcome Score – Extended [IQR] | 8 [2] | 7.5 [1] | 0.49 |
| Overall Cognitive Composite Score mean (SD) | 48.98 (8.66) | 47.48 (7.15) | 0.61 |
| Encephalomalacic lesions present | 73 (39%) | -- | -- |
| <u>Psychological Health</u> |  |  |  |
| RAND-36 emotional wellbeing t score mean (SD) | 50.64 (9.54) | 55.97 (6.70) | 0.12 |
| Neuro QOL depression t score mean (SD) | 46.89 (8.21) | 41.36 (6.10) | 0.048 |
| Neuro QOL anxiety t score mean (SD) | 50.32 (8.34) | 45.73 (5.19) | 0.11 |
| <u>Pre-Injury Psychiatric History (%)</u> |  |  |  |
| *None*<br>*Outpatient consult*<br>*Emergency department*<br>*Single psychiatric hospitalization*<br>*Repeat psychiatric hospitalizations* | 149/161 (93%)<br>5/161 (3%)<br>3/161 (2%)<br>1/161 (1%)<br>3/161 (2%) | 81/92 (88%)<br>5/92 (5%)<br>0/92 (0%)<br>3/92 (3%)<br>3/92 (3%) | 0.22 |
| <u>Pre Injury Suicidality (%)</u> |  |  |  |
| *None*<br>*Thoughts of harm without intent/plan*<br>*Thoughts of harm with intent/plan*<br>*Single suicide attempt*<br>*Multiple suicide attempts* | 138/161 (86%)<br>13/161 (8%)<br>6/161 (4%)<br>1/161 (1%)<br>3/161 (2%) | 73/91 (80%)<br>10/91 (11%)<br>5/91 (6%)<br>3/91 (3%)<br>0/91 (0%) | 0.24 |
| <u>Missing Data</u><br>*MRI*<br>*RAND Emotional wellbeing* | -- | 105/113 (93%)<br>8/113 (7%) | -- |
| *Missingness of certain variables accounts for differences in total N's for some variables. |  |  |  |

### Neurocognitive and Patient-Reported Outcome Measures

All participants underwent a comprehensive battery of neurocognitive tests and completed patient-reported outcome measures. The primary outcome of emotional wellbeing was assessed via the RAND 36-Item Health Survey^32^ emotional wellbeing subscale, derived from 5 questions about mood and wellbeing. This patient-reported measure captures the frequency of positive and negative emotional states (happiness, calmness, nervousness, depression, etc.) over the past four weeks. This score was transformed into a t-score such that a participant with the population mean would have a value of 50 and the population SD would correspond to a change of 10. Higher values correspond to greater emotional wellbeing (i.e., less emotional distress). Secondary outcomes included measures of depression and anxiety derived from the Quality of Life in Neurological Disorders (Neuro-QoL)^33^ short form assessments. We computed the average of all responses on the depression and anxiety short forms (separately) and converted these into t scores in the same manner.

Additionally, we report a summary measure of overall cognitive performance. Overall cognitive performance was measured as a composite score derived from standardized scores of multiple cognitive tests spanning key cognitive domains (memory, language, attention, executive functioning, and processing speed; details provided in Supplementary Methods). Finally, demographic data, as well as information about employment and prior medical and psychiatric history, were obtained by participant report. Beginning in 2020, global level of function was measured with the Glasgow Outcome Scale Extended (GOSE).

### MRI Acquisition and Pre-Processing

Brain MRI scans were acquired at one of two sites on either a Siemens Skyra 3T, Phillips Achieva 3T, or Phillips Ingenia Elition 3T. Sequences included high-resolution T1 multi-echo magnetization-prepared rapid acquisition gradient echo (MEMPRAGE)^34^ and single-shell dMRI. All dMRI scans included two acquisitions with reverse phase-encoding directions. The number of diffusion-encoding directions ranged from 30 to 64, the b-value was 1000 s/mm^2^ for all scans, and the spatial resolution was 2mm isotropic for two scanners, and 1.875 x 1.875 x 2 mm for the third. Complete sequence parameters are provided in Supplementary Table 1.

### Encephalomalacic Cortical Lesion Identification and Labeling

Encephalomalacia (areas of focal cortical tissue loss or cavity/gliosis) were manually identified on T1-weighted images as previously described^14, 35^. The distribution of lesion locations throughout the brain was previously reported^14^. Briefly, encephalomalacic lesions were defined as contiguous areas of disrupted cortex and adjacent white matter not conforming to a vascular distribution. The full extent of the lesion, including any deep white matter involvement, was included as part of the lesion label. As described previously^14^, we registered lesions traced on participant T1 volumes into Montreal Neurological Institute (MNI152) T1 1mm space using linear (rigid + affine) and non-linear (deformable syn) registrations (ANTs)^36^.

### dMRI Pre-processing

The dMRI pre-processing pipeline^37^ included correction for Gibbs ringing^38^ in MRtrix3^39^ and correction for susceptibility induced distortions, motion, and eddy-current distortions in FSL^40, 41^. Motion parameters and contrast-to-noise-ratio maps were estimated to perform data quality control. The diffusion tensor model was fit to the data using DTIFIT in FSL to obtain FA maps. Automated reconstruction of 40 white matter pathways was performed using TRActs Constrained by UnderLying Anatomy (TRACULA)^42, 43^ with default parameters (200 initial iterations and 7,500 sample paths). Estimates of fiber orientation at each voxel were obtained by fitting the ball-and-stick model in FSL using BEDPOSTX^44^. The anatomical segmentations necessary to compute the anatomical priors in TRACULA were obtained from the T1-weighted images using the SamSeg tool in FreeSurfer 7.4.0^45^, and transformed onto the subject’s individual dMRI space using a boundary-based, affine registration method^46^. Participants who failed to complete the TRACULA pipeline due to inaccurate surface reconstructions or poor-quality diffusion data (N=34) were excluded from tract-based analyses.

We measured the weighted (by voxel probability of inclusion in each tract) mean FA within each of the 40 TRACULA-derived volumetric tract labels. We excluded the fornix, as optimal reconstruction requires segmentation of subthalamic nuclei, which is not available in the standard recon-all pipeline^47^. As a summary measure of global white matter integrity, we calculated the mean FA across all reconstructed tracts for each subject. We then converted this mean FA into a standardized Z score. For tract-based analyses, we standardized the FA within each tract.

### Statistical Analysis: Encephalomalacic Lesions

We first tested whether the presence of encephalomalacic lesions (yes/no) was independently associated with emotional wellbeing. We used a generalized linear model, with covariates including age (continuous), sex (M/F), years of education (continuous), time since injury (continuous), and a history of seeking psychiatric care or suicidality (yes/no). Among the subgroup of participants with encephalomalacic lesions, we used univariate linear models to determine whether lesion volume was associated with emotional wellbeing.

To explore the brain-wide distribution of associations between lesion presence and emotional wellbeing, we performed voxel lesion symptom mapping (VLSM). We used Permutation Analysis for Linear Models (PALM) software (FSL) to test voxels lesioned in at least 5 participants. We generated a null distribution using 2,000 random outcome permutations and controlled for multiple comparisons using false discovery rate (FDR) adjustment. To test whether an observed hemispheric discrepancy in positive versus negative associations with emotional wellbeing was greater than would be expected due to chance, we computed the absolute difference in the sum of T values in each hemisphere. We built a null distribution for this value from the maps generated from each of the 2,000 outcome permutations and used that distribution to report an empirical p value for the observed difference.

### Statistical Analysis: White matter Integrity

We used multivariate linear model to test the association between global mean white matter integrity (FA) and emotional well-being, adjusting for age, sex, years of education, time since injury, history of seeking psychiatric care or suicidality, scanner, contrast-to-noise ratio of the non-diffusion-weighted (i.e., b0) volume from the dMRI scan, and absolute motion.

In a data-driven analysis intended to identify the strongest regional associations within the white matter, we fit separate models (using the same covariates as above) with the standardized mean FA in each of 40 TRACULA-reconstructed white matter tracts. We performed a Holm adjustment of the resulting coefficient p values to control the family wise error rate.

All analyses were performed using R (RStudio2023.06.2).

## Results

### Participant Characteristics

We analyzed participants in the LETBI cohort with chronic TBI who underwent MRI and behavioral assessment (N=188; Table 1; Figure 1). The mean age was 58 (SD 15), 129 (68%) were male, 166 (88%) were white, and the median years since the TBI was 8 (IQR 14 years). Seventy-three (39%) participants had encephalomalacic cortical lesions visible on T1 MRI, most prominently in the inferomedial frontal and inferolateral temporal lobes (Supplementary Figure 1). The mean RAND emotional wellbeing t score was near the population mean (50.7 [SD 9.5]; Table 1). Participants also scored near the population mean for the Neuro-QoL Depression (46.9 [8.2]; Table 1), and Neuro-QoL Anxiety t scores (50.3 [8.3]; Table 1). The mean overall cognitive composite t score was 49.0 (SD 8.6; Table 1).

**Figure 1:**
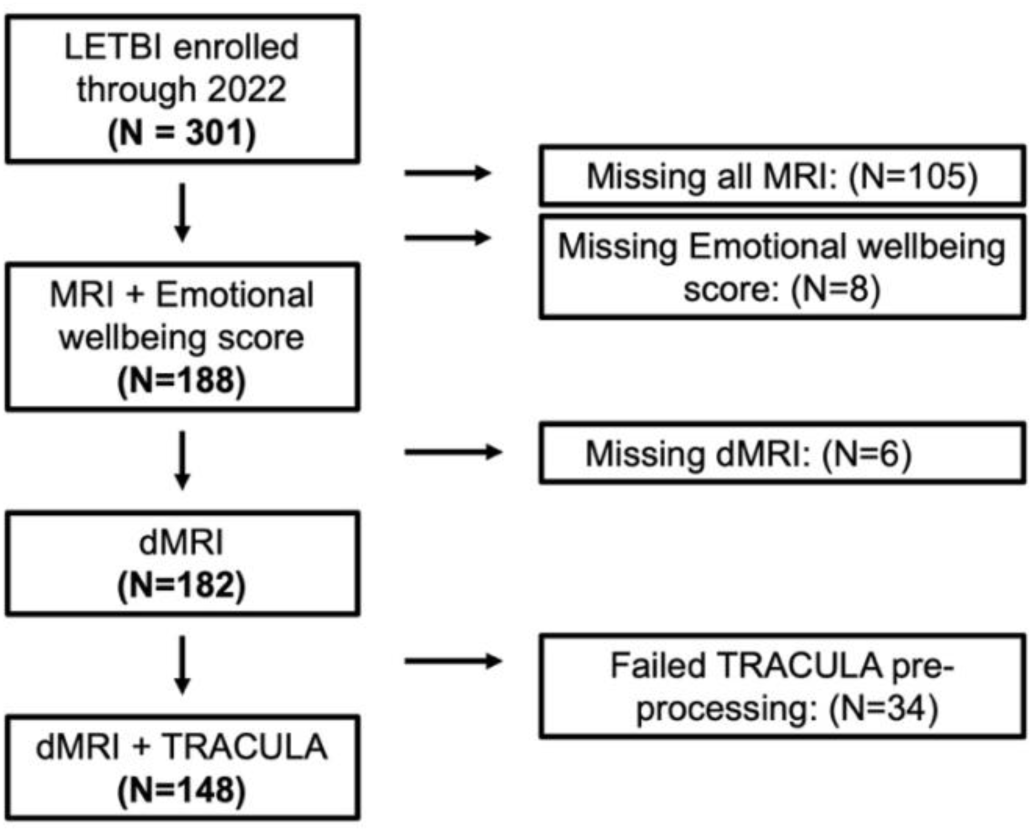
Study CONSORT diagram. Abbreviations: LETBI Late Effects of Traumatic Brain Injury; TRACULA Tracts Constrained by Underlying Anatomy

Compared to the rest of the cohort, participants in the bottom quartile of emotional wellbeing had similar cognitive performance, but were younger (p=0.041) and more likely to have reported being disabled for their employment status (p=0.002; Supplementary Table 2).

### Encephalomalacic Lesions and Emotional Wellbeing

The presence of any encephalomalacic lesion was independently associated with higher emotional wellbeing scores (4.1 [1.0, 7.0] point *increase* in RAND emotional wellbeing t score; p=0.009; Table 2). Fifteen percent of participants with lesions scored in the lowest emotional wellbeing quartile compared to 31% of those without lesions. Overall level of function was no different between participants with (median GOSE 8, SD [2.8]) or without (7 [2]; p=0.81) lesions.

**Table 2:**
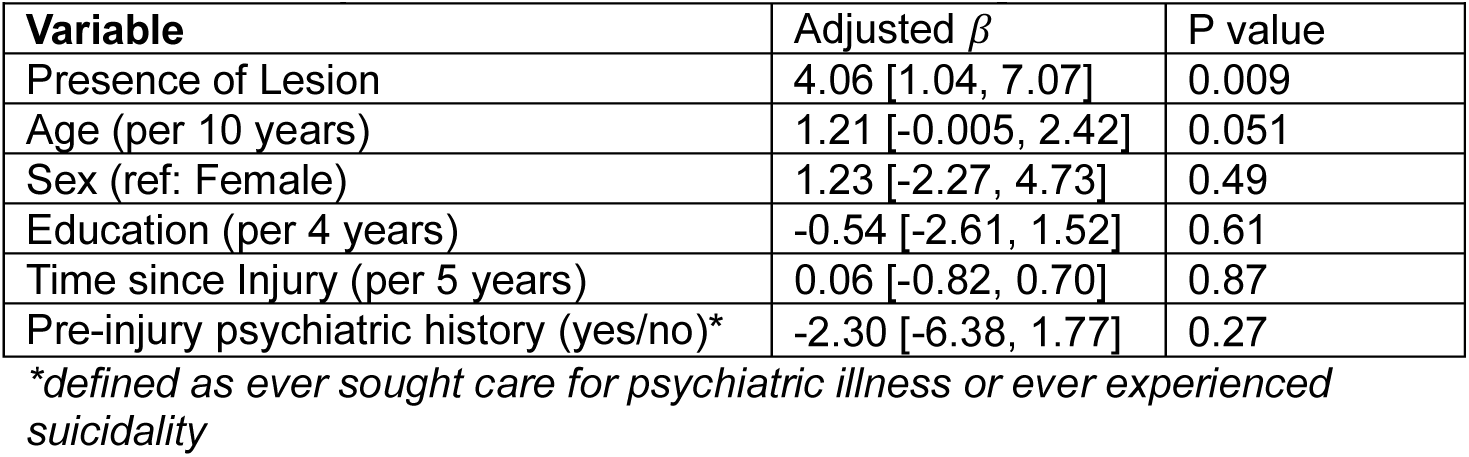
Lesion presence and emotional wellbeing.

The presence of lesions remained associated with better emotional wellbeing after also controlling for overall cognition (4.6 [1.0, 8.2]; p = 0.01; Supplementary Table 3), and when using secondary measures of anxiety (3.8 [6.4, 1.1] point *decrease* [improvement] in anxiety t score; p=0.006; Supplementary Table 4) and depression t score (4.1 [6.7, 1.5] point *decrease* [improvement] in depression score; p=0.003; Supplementary Table 5).

Among participants with lesions, there was no association between lesion size and emotional wellbeing (0.03 [-0.04, 0.1] point change per cc^3^ of lesion volume p=0.47; Supplementary Table 6). In the data-driven, voxel-wise analysis, right temporal and lateral orbitofrontal lesions were associated with lower (worse) emotional wellbeing scores, and left hemispheric lesions associated with higher (better) emotional wellbeing scores (Figure 2A). A hemispheric discrepancy in statistical values at least as extreme was unlikely to be due to chance alone (Ppermutation = 0.026). Though the strongest association was observed in the right inferior temporal lobe—where lesion presence was associated with *lower* emotional wellbeing scores (PFDR < 0.05; Figure 2B)—most encephalomalacic lesions overlapped areas associated with higher emotional wellbeing scores (Supplementary Figure 2).

**Figure 2:**
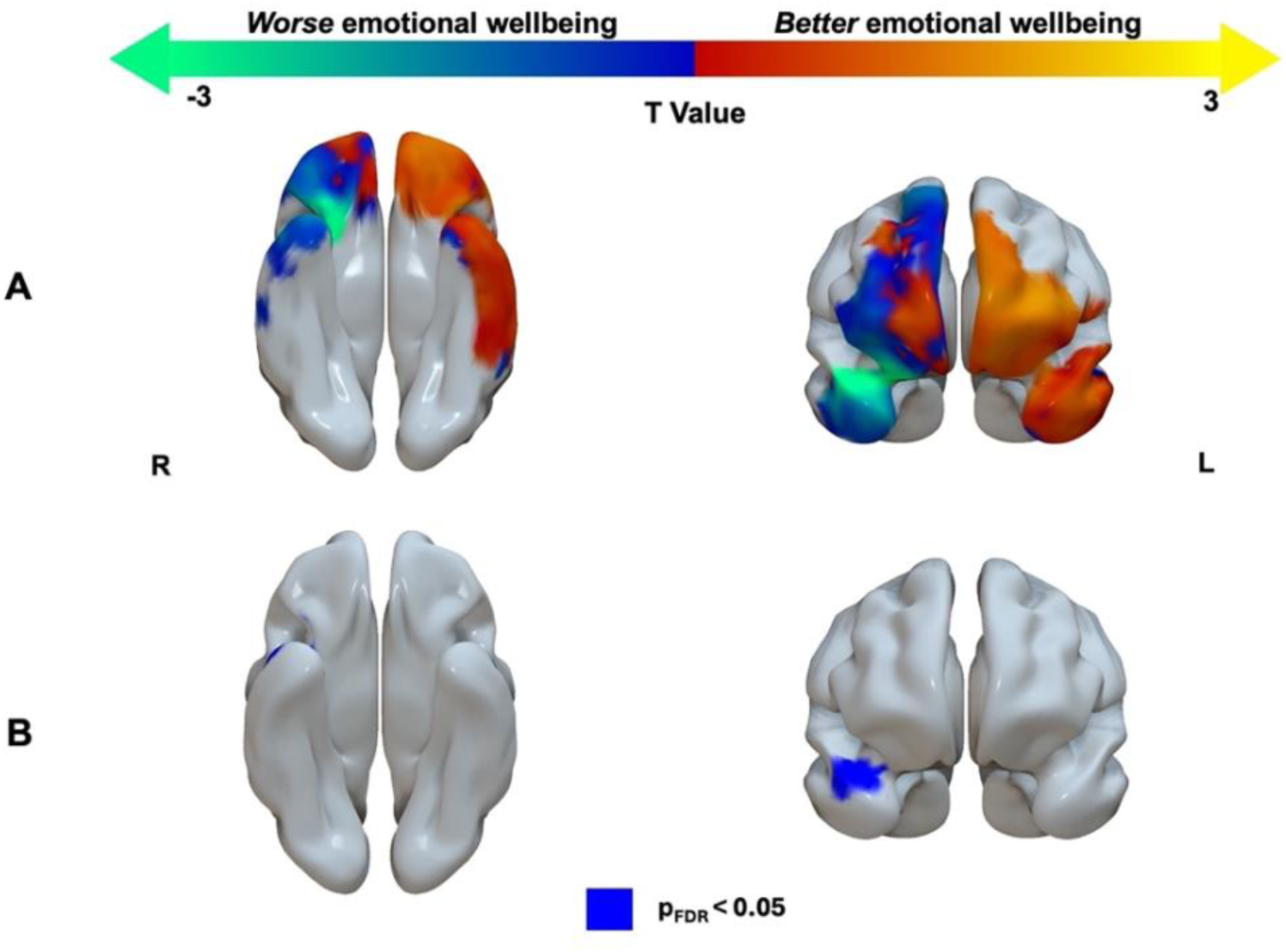
Cortical lesion locations associated with emotional wellbeing. (A) Results from voxel lesion symptom mapping analysis including cortical voxels lesioned in at least 5 participants. The un-thresholded statistical T map is shown. (B) A single cluster in the R temporal lobe had FDR-adjusted p values < 0.05.

### White Matter Integrity and Emotional Wellbeing

Globally reduced white matter integrity was independently associated with better emotional wellbeing (2.7 [0.8, 4.7] same point *increase* in RAND emotional wellbeing t score per 1 SD *decrease* in mean FA; p=0.008; Table 3). This finding persisted when using secondary measures of anxiety (2.7 [1.0, 4.5] point *decrease* [improvement] in anxiety t score; p =0.002; Supplementary Table 7) and depression t score (2.3 [0.6, 4.0] point *decrease* [improvement] in depression score; p = 0.003; Supplementary Table 8).

**Table 3:** Mean white matter integrity and emotional wellbeing.

| **Table 3: Mean white matter integrity and emotional wellbeing** |  |  |
| --- | --- | --- |
| **Variable** | **Adjusted $\beta$** | **P value** |
| Mean white matter FA | -2.71 [-4.67, -0.76] | 0.008 |
| Age (per 10 years) | 0.42 [-1.01, 1.85] | 0.57 |
| Sex (ref. Female) | -0.88 [-4.72, 2.97] | 0.66 |
| Education (per 4 years) | -1.50 [-3.77, 0.78] | 0.20 |
| Time since Injury (per 5 years) | 0.09 [-0.70, 0.88] | 0.83 |
| **Scanner** (ref: Ingenia) |  |  |
| Skyra | -6.90 [-16.13, 2.32] | 0.15 |
| CNR* | 2.27 [-9.00, 13.54] | 0.69 |
| Absolute Motion (mm) | -4.57 [-9.45, 0.31] | 0.07 |
| Pre-injury psychiatric history (yes/no)** | -3.75 [-8.53, 1.03] | 0.13 |

\*Contrast to noise ratio
\*\*defined as ever sought care for psychiatric illness or ever experienced suicidality

The strongest associations between individual white matter tract integrity and emotional wellbeing were in the anterior (genu) corpus callosum (Figure 3; β = -3.0, [-1.3, -4.7]; pHolm=0.034) and the left cingulum bundle (Figure 3; β = -3.0, [-1.2, -4.8]; pHolm=0.048).

**Figure 3:**
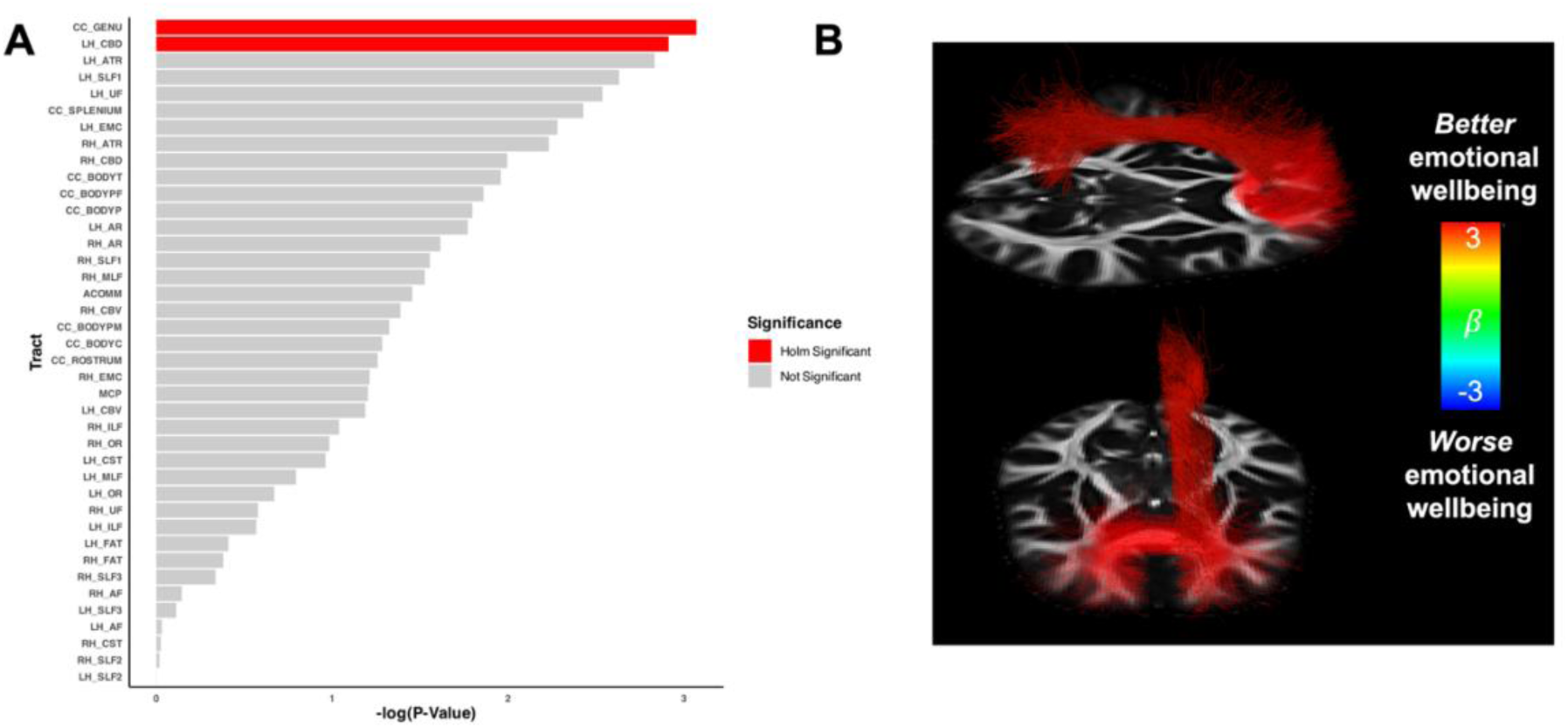
Individual white matter tract associations with emotional wellbeing. (A) Waterfall plot of p values representing the negative log of the uncorrected p values associated with the coefficient in a fully adjusted regression between the mean FA in each tract and emotional wellbeing. Red bars have a Holm (FWE) adjusted p value < 0.05. (B) Hom-significant tracts shown on the TRACULA template brain from diffusion data averaged from 35 Human Connectome Project participants (Maffei et al. *NeuroImage* 2021). The color corresponds to the regression coefficient quantifying the adjusted association with emotional wellbeing. Abbreviations: ACOMM: anterior commissure; AF: arcuate fasciculus; AR: acoustic radiation; ATR: anterior thalamic radiation; CC BODYC: central section of the body of the CC; CC BODYP: parietal section of the body of the CC; CC BODYPF: prefrontal section of the body of the CC; CC BODYPM: premotor section of the body of the CC; CC BODYT: temporal section of the body of the CC; CC GENU: genu of the CC; CC ROSTRUM: rostrum of the CC; CC SPLENIUM: splenium of the CC; CBD: dorsal portion of cingulum bundle; CBV: ventral portion of the cingulum bundle; CC: corpus callosum; CST: cortico-spinal tract; EMC: extreme capsule; FAT: frontal Aslant tract; ILF: inferior longitudinal fasciculus; LH: left hemisphere; ILF: middle longitudinal fasciculus; OR: optic radiation; RH: right hemisphere; SLF I,II,III: first, second, and third branch of the superior longitudinal fasciculus; UF: uncinate fasciculus.

## Discussion

Contrary to prior studies in chronic TBI^20, 30, 48^, we found that a greater burden of structural brain injury was paradoxically associated with *better* self-reported emotional wellbeing. Both encephalomalacic lesions and reduced white matter integrity were each associated with *greater* emotional wellbeing and *lower* scores for depression and anxiety. These findings prompt a critical re-appraisal of our understanding of emotional wellbeing after TBI.

While some studies have suggested that emotional disturbances are more prevalent after milder TBIs (suggested by ancillary markers of injury severity)^5, 11, 13^, others have found the opposite—with the incidence of depression after moderate/severe TBI^7^ exceeding the incidence after mild TBI^4, 8^. In our study, participants with encephalomalacic lesions were half as likely to score in the lowest emotional well being quartile, as compared to those without lesions.

Recognizing that prior studies have shown that psychological resilience, coping strategies, and premorbid traits influence long-term emotional outcomes^5, 8^, we accounted for multiple potential confounding factors and found that the associations reported here remained after accounting for age, a history of psychiatric hospitalization and overall cognitive performance. In theory, these findings could be explained by the disability paradox, wherein some patients with high degrees of disability and dependency also report high satisfaction with life^49^. Three pieces of evidence from our study argue against this. First, the association with emotional wellbeing was independent of overall cognition. If severely disabled participants reported better emotional wellbeing, then adjusting for overall cognition should at least partially address this potential confounding. Second, participants in the bottom emotional wellbeing quartile were *more,* not *less* likely to report disability as their employment status. Finally, GOSE scores, which reflect overall level of function, were not found to systematically differ among participants with verus without lesions.

One plausible explanation for this finding would be a protective effect of specific types of structural injury against emotional disturbances. Indeed, a seminal study of Vietnam War veterans with TBI found that post-traumatic stress disorder was less common in participants with structural brain lesions^50^. The majority of the lesions in this study were situated inferiorly and ventrally to structures traditionally associated with depressive symptoms, such as the dlPFC and sgACC^25, 26, 51^. A previous study of ventromedial frontal lesions in patients with penetrating TBI also found these lesion locations to be associated with fewer symptoms of depression^52^. Given that only 73 participants in our study had encephalomalacic lesions, we likely had less power to detect regional associations than prior lesion-based studies not restricted to individuals with TBI^25^.

In spite of this limited power, we identified a striking left/right dissociation in the direction of the association between lesions and emotional wellbeing—with left-sided lesions generally associated with better emotional wellbeing and right-sided (inferomedial frontal and inferolateral temporal) lesions generally associated with worse emotional wellbeing. Indeed, the strongest brain-wide signal was in the direction of worse emotional wellbeing with right temporal lobe lesions. Prior studies of patients with temporal lobe epilepsy before and after surgery have suggested worsening mood after right temporal resections^53–56^, but the totality of evidence appears inconsistent^57, 58^.

Systematic reviews in ischemic stroke have found no consistent effect of lesion hemisphere on depressive symptoms^59^. Studies using the more modern technique of lesion network mapping^24^ have implicated a brain-wide network of regions most prominently defined by the sgACC and dlPFC^25, 26^. In this network^25, 26^, lesions of either temporal lobe are associated with better emotional wellbeing. This regional effect differs from what we observed here, where the direction of association in the temporal lobe varied by hemisphere. It is possible that the neurophysiology of mood disorders after TBI may have an entirely different neurobiological basis than mood disorders that occur independently of TBI^60^. Such a dissociation is supported by functional brain mapping studies in depressed patients with TBI^29^.

Lower white matter integrity was also associated with better emotional wellbeing, with the strongest associations observed in the genu of the corpus callosum and the cingulum bundle. These tracts contain fibers emanating from subgenual anterior cingulate cortex^61, 62^. Traditional neurophysiological models of depression implicate aberrantly high sgACC activity^63, 64^ and functional connectivity^65^. Reduced integrity of the sgACC’s efferent white matter connections may functionally disconnect it, limiting susceptibility to mood disturbances. The tract with the third strongest association, falling just outside the adjusted significance threshold, was the left anterior thalamic radiation. This tract has been shown to be a key mediator of functional connectivity changes in patients who respond to transcranial magnetic stimulation treatment for depression^66^.

It is important to emphasize that the effect size for the association between white matter integrity and emotional wellbeing was small. A 1 SD reduction in global white matter FA was associated with a roughly 2.5-point increase in the emotional wellbeing t score (25% of the population SD). A 1 SD increase in emotional wellbeing would therefore only be seen in someone with white matter integrity approximately 4 SDs below the study mean. This observation aligns with prior studies indicating that while dMRI can detect microstructural injury relevant to cognition, many other factors—including premorbid abilities, psychosocial circumstances, and compensatory neuroplasticity— may influence post-TBI cognitive function^67^.

Prior neuroiomaging investigations have found that brain microstructural abnormalities are more prevalent in chronic TBI patients with emotional disturbances^20, 30, 48^. One such study in a multi-center US cohort reported a negative association between some dMRI measures of white matter microstructural integrity and emotional resilience^30^—the opposite direction of the findings reported here. Differences in study design may explain the discrepant findings, which most prominently include a different outcome measure (resilience [encompassing concomitant cognitive impairment] versus wellbeing), subacute versus chronic assessment time points, distinct dMRI-measured microstructural metrics, and absence versus presence of covariate adjustment.

It is also important to consider that the LETBI study was designed to enroll individuals with chronic TBI from the community. This community-based enrollment should be considered when interpreting the observed relationship between structural injury and emotional wellbeing, as the current sample may better represent the population of those living with TBI relative to prior studies that enrolled individuals exclusively from specialized clinics.

### Limitations

These results should be interpreted in the context of several limitations. First, the long delays between injury and behavioral assessment (i.e., average time post-injury at time of assessment Mean [SD] ) limit the ability to infer causality in cross-sectional data. Longitudinal assessments, which are currently being conducted as part of the ongoing LETBI study, are needed to better clarify temporal relationships. Second, enrollment bias may have systematically excluded individuals with severe impairments and severe structural injuries. Third, the primary emotional outcome was a self-reported summary measure of emotional wellbeing, which does not capture irritability, disinhibition, or alexithymia. Fourth, given the limited number of participants with lesions, analyses of lesion location were likely underpowered. Fifth, the diffusion MRI data were acquired across several scanners. While our models included scanner as a covariate, we lacked a matched control sample to perform full cross-site harmonization of the dMRI scans. Finally, given our lack of control of the study-level alpha, there is a risk of false positive findings, and these results should be viewed as primarily hypothesis-generating.

### Conclusions

In a cohort of 188 individuals with chronic TBI from the LETBI study, we found that worse structural injury was associated with better emotional wellbeing. These findings challenge prior assumptions about the neuroanatomic basis of emotional disturbances after TBI. Longitudinal studies are needed to establish the causal nature of this relationship and further elucidate the responsible brain-wide networks.

## Data Availability

All data produced in the present study are available upon reasonable request to the authors

## Acknowledgements

We thank the LETBI study participants and their families.

## References

1. Ponsford JL, Downing MG, Olver J, et al. Longitudinal follow-up of patients with traumatic brain injury: outcome at two, five, and ten years post-injury. J Neurotrauma 2014;31:64–77.

2. Konrad C, Geburek AJ, Rist F, et al. Long-term cognitive and emotional consequences of mild traumatic brain injury. Psychol Med 2011;41:1197–1211.

3. Engberg AW, Teasdale TW. Psychosocial outcome following traumatic brain injury in adults: a long-term population-based follow-up. Brain Inj 2004;18:533–545.

4. Howlett JR, Nelson LD, Stein MB. Mental Health Consequences of Traumatic Brain Injury. Biol Psychiatry 2022;91:413–420.

5. Brett BL, Kramer MD, Whyte J, et al. Latent Profile Analysis of Neuropsychiatric Symptoms and Cognitive Function of Adults 2 Weeks After Traumatic Brain Injury: Findings From the TRACK-TBI Study. JAMA Netw Open 2021;4:e213467.

6. Kreutzer JS, Seel RT, Gourley E. The prevalence and symptom rates of depression after traumatic brain injury: a comprehensive examination. Brain Inj 2001;15:563–576.

7. Bombardier CH, Fann JR, Temkin NR, Esselman PC, Barber J, Dikmen SS. Rates of major depressive disorder and clinical outcomes following traumatic brain injury. JAMA 2010;303:1938–1945.

8. Stein MB, Jain S, Giacino JT, et al. Risk of Posttraumatic Stress Disorder and Major Depression in Civilian Patients After Mild Traumatic Brain Injury: A TRACK-TBI Study. JAMA Psychiatry 2019;76:249–258.

9. Valk-Kleibeuker L, Heijenbrok-Kal MH, Ribbers GM. Mood after moderate and severe traumatic brain injury: a prospective cohort study. PLoS One 2014;9:e87414.

10. Malec JF, Brown AW, Moessner AM, Stump TE, Monahan P. A preliminary model for posttraumatic brain injury depression. Arch Phys Med Rehabil 2010;91:1087–1097.

11. Nelson LD, Kramer MD, Joyner KJ, et al. Relationship between transdiagnostic dimensions of psychopathology and traumatic brain injury (TBI): A TRACK-TBI study. J Abnorm Psychol 2021;130:423–434.

12. Schonberger M, Ponsford J, Gould KR, Johnston L. The temporal relationship between depression, anxiety, and functional status after traumatic brain injury: a cross-lagged analysis. J Int Neuropsychol Soc 2011;17:781–787.

13. Choi Y, Kim EY, Sun J, et al. Incidence of Depression after Traumatic Brain Injury: A Nationwide Longitudinal Study of 2.2 Million Adults. J Neurotrauma 2022;39:390–397.

14. Snider SB, Gilmore N, Freeman HJ, et al. Cortical lesions and focal white matter injury are associated with attentional performance in chronic traumatic brain injury. Brain Commun 2025;7:fcae420.

15. Diamond BR, Donald CLM, Frau-Pascual A, et al. Optimizing the accuracy of cortical volumetric analysis in traumatic brain injury. MethodsX 2020;7:100994.

16. Kraus MF, Susmaras T, Caughlin BP, Walker CJ, Sweeney JA, Little DM. White matter integrity and cognition in chronic traumatic brain injury: a diffusion tensor imaging study. Brain 2007;130:2508–2519.

17. Kinnunen KM, Greenwood R, Powell JH, et al. White matter damage and cognitive impairment after traumatic brain injury. Brain 2011;134:449–463.

18. Snider SB, Bodien YG, Bianciardi M, Brown EN, Wu O, Edlow BL. Disruption of the ascending arousal network in acute traumatic disorders of consciousness. Neurology 2019;93:e1281–e1287.

19. Jolly AE, Balaet M, Azor A, et al. Detecting axonal injury in individual patients after traumatic brain injury. Brain 2021;144:92–113.

20. Medeiros GC, Twose C, Weller A, et al. Neuroimaging Correlates of Depression after Traumatic Brain Injury: A Systematic Review. J Neurotrauma 2022;39:755–772.

21. Bender Pape TL, Herrold AA, Guernon A, Aaronson A, Rosenow JM. Neuromodulatory Interventions for Traumatic Brain Injury. J Head Trauma Rehabil 2020;35:365–370.

22. Shin SS, Dixon CE, Okonkwo DO, Richardson RM. Neurostimulation for traumatic brain injury. J Neurosurg 2014;121:1219–1231.

23. Kahana MJ, Ezzyat Y, Wanda PA, et al. Biomarker-guided neuromodulation aids memory in traumatic brain injury. Brain Stimul 2023;16:1086–1093.

24. Fox MD. Mapping Symptoms to Brain Networks with the Human Connectome. N Engl J Med 2018;379:2237–2245.

25. Siddiqi SH, Schaper F, Horn A, et al. Brain stimulation and brain lesions converge on common causal circuits in neuropsychiatric disease. Nat Hum Behav 2021;5:1707–1716.

26. Padmanabhan JL, Cooke D, Joutsa J, et al. A Human Depression Circuit Derived From Focal Brain Lesions. Biol Psychiatry 2019;86:749–758.

27. Siddiqi SH, Taylor SF, Cooke D, Pascual-Leone A, George MS, Fox MD. Distinct Symptom-Specific Treatment Targets for Circuit-Based Neuromodulation. Am J Psychiatry 2020;177:435–446.

28. Siddiqi SH, Philip NS, Palm ST, et al. A potential target for noninvasive neuromodulation of PTSD symptoms derived from focal brain lesions in veterans. Nat Neurosci 2024;27:2231–2239.

29. Siddiqi SH, Kandala S, Hacker CD, et al. Precision functional MRI mapping reveals distinct connectivity patterns for depression associated with traumatic brain injury. Sci Transl Med 2023;15:eabn0441.

30. Cai LT, Brett BL, Palacios EM, et al. Emotional Resilience Predicts Preserved White Matter Microstructure Following Mild Traumatic Brain Injury. Biol Psychiatry Cogn Neurosci Neuroimaging 2024;9:164–175.

31. Edlow BL, Keene CD, Perl DP, et al. Multimodal Characterization of the Late Effects of Traumatic Brain Injury: A Methodological Overview of the Late Effects of Traumatic Brain Injury Project. J Neurotrauma 2018;35:1604–1619.

32. Hays RD, Sherbourne CD, Mazel RM. The RAND 36-Item Health Survey 1.0. Health Econ 1993;2:217–227.

33. Cella D, Lai JS, Nowinski CJ, et al. Neuro-QOL: brief measures of health-related quality of life for clinical research in neurology. Neurology 2012;78:1860–1867.

34. van der Kouwe AJW, Benner T, Salat DH, Fischl B. Brain morphometry with multiecho MPRAGE. NeuroImage 2008;40:559–569.

35. Freeman HJ, Atalay AS, Li J, et al. Cortical Lesion Expansion in Chronic Traumatic Brain Injury. medRxiv 2025; DOI 0.1101/2024.06.24.24309307.

36. Tustison NJ, Cook PA, Holbrook AJ, et al. The ANTsX ecosystem for quantitative biological and medical imaging. Sci Rep 2021;11:9068.

37. Veraart J, Novikov DS, Christiaens D, Ades-Aron B, Sijbers J, Fieremans E. Denoising of diffusion MRI using random matrix theory. Neuroimage 2016;142:394–406.

38. Kellner E, Dhital B, Kiselev VG, Reisert M. Gibbs-ringing artifact removal based on local subvoxel-shifts. Magn Reson Med 2016;76:1574–1581.

39. Tournier JD, Smith R, Raffelt D, et al. MRtrix3: A fast, flexible and open software framework for medical image processing and visualisation. Neuroimage 2019;202:116137.

40. Andersson JLR, Sotiropoulos SN. An integrated approach to correction for off-resonance effects and subject movement in diffusion MR imaging. Neuroimage 2016;125:1063–1078.

41. Andersson JL, Skare S, Ashburner J. How to correct susceptibility distortions in spin-echo echo-planar images: application to diffusion tensor imaging. Neuroimage 2003;20:870–888.

42. Yendiki A, Panneck P, Srinivasan P, et al. Automated probabilistic reconstruction of white-matter pathways in health and disease using an atlas of the underlying anatomy. Front Neuroinform 2011;5:23.

43. Maffei C, Lee C, Planich M, et al. Using diffusion MRI data acquired with ultra-high gradient strength to improve tractography in routine-quality data. Neuroimage 2021;245:118706.

44. Behrens TE, Johansen-Berg H, Woolrich MW, et al. Non-invasive mapping of connections between human thalamus and cortex using diffusion imaging. Nat Neurosci 2003;6:750–757.

45. Fischl B, van der Kouwe A, Destrieux C, et al. Automatically parcellating the human cerebral cortex. Cereb Cortex 2004;14:11–22.

46. Greve DN, Fischl B. Accurate and robust brain image alignment using boundary-based registration. Neuroimage 2009;48:63–72.

47. Maffei C, Gilmore N, Snider SB, et al. Automated detection of axonal damage along white matter tracts in acute severe traumatic brain injury. Neuroimage Clin 2023;37:103294.

48. Schmidt AT, Lindsey HM, Dennis E, et al. Diffusion Tensor Imaging Correlates of Resilience Following Adolescent Traumatic Brain Injury. Cogn Behav Neurol 2021;34:259–274.

49. Albrecht GL, Devlieger PJ. The disability paradox: high quality of life against all odds. Soc Sci Med 1999;48:977–988.

50. Koenigs M, Huey ED, Raymont V, et al. Focal brain damage protects against post-traumatic stress disorder in combat veterans. Nat Neurosci 2008;11:232–237.

51. Mayberg HS. Defining the neural circuitry of depression: toward a new nosology with therapeutic implications. Biol Psychiatry 2007;61:729–730.

52. Koenigs M, Huey ED, Calamia M, Raymont V, Tranel D, Grafman J. Distinct regions of prefrontal cortex mediate resistance and vulnerability to depression. J Neurosci 2008;28:12341–12348.

53. Irle E, Peper M, Wowra B, Kunze S. Mood changes after surgery for tumors of the cerebral cortex. Arch Neurol 1994;51:164–174.

54. Kohler C, Norstrand JA, Baltuch G, et al. Depression in temporal lobe epilepsy before epilepsy surgery. Epilepsia 1999;40:336–340.

55. Quigg M, Broshek DK, Heidal-Schiltz S, Maedgen JW, Bertram EH, 3rd. Depression in intractable partial epilepsy varies by laterality of focus and surgery. Epilepsia 2003;44:419–424.

56. Glosser G, Zwil AS, Glosser DS, O’Connor MJ, Sperling MR. Psychiatric aspects of temporal lobe epilepsy before and after anterior temporal lobectomy. J Neurol Neurosurg Psychiatry 2000;68:53–58.

57. Malmgren K, Starmark JE, Ekstedt G, Rosen H, Sjoberg-Larsson C. Nonorganic and Organic Psychiatric Disorders in Patients after Epilepsy Surgery. Epilepsy Behav 2002;3:67–75.

58. Meldolesi GN, Di Gennaro G, Quarato PP, et al. Changes in depression, anxiety, anger, and personality after resective surgery for drug-resistant temporal lobe epilepsy: a 2-year follow-up study. Epilepsy Res 2007;77:22–30.

59. Carson AJ, MacHale S, Allen K, et al. Depression after stroke and lesion location: a systematic review. Lancet 2000;356:122–126.

60. Li LM, Carson A, Dams-O’Connor K. Psychiatric sequelae of traumatic brain injury - future directions in research. Nat Rev Neurol 2023;19:556–571.

61. Shi W, Xue M, Wu F, et al. Whole-brain mapping of efferent projections of the anterior cingulate cortex in adult male mice. Mol Pain 2022;18:17448069221094529.

62. Tsang P, Cheng FC. Recent advances in the local treatment of thermal injury. Xianggang Hu Li Za Zhi 1978:21–26.

63. Mayberg HS, Lozano AM, Voon V, et al. Deep brain stimulation for treatment-resistant depression. Neuron 2005;45:651–660.

64. Mayberg HS, Brannan SK, Tekell JL, et al. Regional metabolic effects of fluoxetine in major depression: serial changes and relationship to clinical response. Biol Psychiatry 2000;48:830–843.

65. Connolly CG, Wu J, Ho TC, et al. Resting-state functional connectivity of subgenual anterior cingulate cortex in depressed adolescents. Biol Psychiatry 2013;74:898–907.

66. Barredo J, Bellone JA, Edwards M, Carpenter LL, Correia S, Philip NS. White matter integrity and functional predictors of response to repetitive transcranial magnetic stimulation for posttraumatic stress disorder and major depression. Depress Anxiety 2019;36:1047–1057.

67. Niogi SN, Mukherjee P. Diffusion tensor imaging of mild traumatic brain injury. J Head Trauma Rehabil 2010;25:241–255.

