## Supplementary Material for "Structural injuries are associated with better emotional wellbeing after traumatic brain injury"

Samuel B. Snider^1^, Hui Shi^1^, Chiara Maffei^2,3,4^, Natalie Gilmore^3,5,6^, Holly J. Freeman^2,3^, Alexander S. Atalay^2,3^, Jian Li^2,3,4^, Jessica A. Yeager^2,3^, Raj G. Kumar^7^, Belinda Yew^7^, Erin M. Conley^7^, Ariana L. Velazquez^7^, Julia Kirschenbaum^7^, Enna Selmanovic^7^, Holly Carrington^7^, Yelena G. Bodien^2,3,8^, Jeanne M. Hoffman^9^, Christine Mac Donald^10^, Brian L. Edlow^2,3,4^*, Kristen Dams-O'Connor^7,11^*

* co-senior authors

**Affiliations:**

1 Department of Neurology, Brigham and Women’s Hospital and Harvard Medical School, Boston MA, USA

2 Center for Neurotechnology and Neurorecovery, Massachusetts General Hospital, Boston, MA, USA

3 Department of Neurology, Massachusetts General Hospital and Harvard Medical School, Boston, MA, USA

4 Athinoula A. Martinos Center for Biomedical Imaging, Department of Radiology, Massachusetts General Hospital and Harvard Medical School, Charlestown, MA, USA

5 Tampa VA Research and Education Foundation, Tampa, VA, USA

6 Research Service, James A. Haley Veterans’ Hospital, Tampa, VA, USA

7 Department of Rehabilitation and Human Performance, Icahn School of Medicine at Mount Sinai, New York, NY, USA

8 Department of Surgery, Department of Physical Medicine & Rehabilitation, Department of Neurological Surgery, Vanderbilt University Medical Center, Nashville TN USA

9 Department of Rehabilitation Medicine, University of Washington School of Medicine, Seattle, WA

10 Department of Neurological Surgery, University of Washington, Seattle, WA, USA

11 Department of Neurology, Icahn School of Medicine at Mount Sinai, New York, NY, USA

**Contents:**

Supplementary Methods: Page 2

Supplementary Tables 1-8: Pages 3-7

Supplementary Figures: Page 8

References: Page 9

**Supplementary Methods:**

*Overall Cognitive Performance*

Relative to individual measures, composite scores of cognitive performance have been shown to have stronger associations with neuropathology markers in cohorts with neurodegenerative disorders^1^. Overall cognitive performance was computed used population-normalized t scores from select tests (California Verbal Learning Test 2 long-delay free recall^2^ t score [normalized by age, sex, and education]; mean 45.93 SD [14.38], Digit Span t score [normalized by age]; mean 50.22 [10.05], Wechsler Adult Intelligence Scale Symbol Search^3^ t score [normalized by age]; mean 48.29 [11.24], and the Controlled Oral Word Association Test^4^ [COWAT] t score [normalized by age, sex, and education]; mean 48.24 [11.80]). In the general population, t scores are scaled to have a mean of 50 and a standard deviation (SD) of 10. For each measure, higher scores indicate better performance. Overall cognitive performance was defined as the mean of the above four t scores.

| **Supplementary Table 1:** Diffusion MRI acquisition parameters | | | |
| --- | --- | --- | --- |
|  | **Skyra** | **Achieva** | **Ingenia** |
| Diffusion directions | 30 | 32 | 64 |
| B_0_ volumes | 1 | 1 | 9 |
| Voxel size | 2mm isotropic | 2mm isotropic | 1.875x1.875x2.0mm |
| Repetition time | 7 seconds | 7.85 seconds | 3.6 seconds |
| Echo time | 0.089 seconds | 0.07 seconds | 0.084 seconds |
| Phase encode directions | 2 | 2 | 2 |

| **Supplementary Table 2:** Characteristics by emotional wellbeing quartile | | | |
| --- | --- | --- | --- |
|  | Rand Emotional Wellbeing 1^st^ Quartile  ($\leq$43.5; N=47) | Rand Emotional Wellbeing (all others;  N=141) | Difference P Value |
| Mean Age (SD) | 54 (13) | 59 (15) | 0.041 |
| Male Sex (%) | 30/47 (64%) | 99/141 (70%) | 0.53 |
| White Race (%) | 41/47 (87%) | 125/141 (89%) | 0.82 |
| Attended College (%) | 34/47 (72%) | 94/141 (67%) | 0.59 |
| Marital Status (%)  *Never married*  *Married/partnered*  *Divorced/widowed* | 16/47 (34%)  16/47 (34%)  15/47 (32%) | 29/141 (21%)  67/141 (48%)  45/141 (32%) | 0.13 |
| Employment (%)  *Working/Student*  *Unemployed*  *Retired*  *Disabled*  *Other* | 10/47 (21%)  3/47 (6%)  9/47 (19%)  21/47 (45%)  4/47 (9%) | 53/141 (38%)  7/141 (5%)  47/141 (33%)  24/141 (17%)  10/141 (7%) | 0.002 |
| Median years since most recent TBI (IQR) | 8 (14) | 8 (15) | 0.38 |
| Lesions Present | 11/47 (23%) | 62/141 (44%) | 0.02 |
| DTI Tracula Mean Scale (SD) | 0.08 (1.13) | -0.05 (0.97) | 0.47 |
| Rand Emotional Wellbeing t Score mean (SD) | 36.95 (4.78) | 55.21 (5.48) | <0.001 |
| CVLT long delay free recall t mean (SD) | 45.78 (17.42) | 45.98 (13.17) | 0.94 |
| Symbol Search t Score mean (SD) | 46.38 (10.82) | 48.91 (11.34) | 0.19 |
| Digit Span t Score mean (SD) | 49.15 (10.17) | 50.60 (10.02) | 0.40 |
| COWAT Words t Score mean (SD) | 46.48 (13.76) | 48.82 (11.09) | 0.27 |
| Overall Cognitive Composite Score mean (SD) | 47.71 (9.06) | 49.49 (8.49) | 0.27 |
| Mean QOL Depression t score (SD) | 55.84 (6.17) | 43.84 (6.41) | <0.001 |
| Mean QOL Anxiety t Score (SD) | 58.68 (6.04) | 47.49 (7.01) | <0.001 |
| Pre Injury Psychiatric History (%)  *None*  *Have sought outpatient consult*  *Have gone to ED*  *Single psychiatric hospitalization*  *Repeat psychiatric hospitalizations* | 39 (89%)  3 (7%)  1 (2%)  0 (0%)  1 (2%) | 110 (94%)  2 (2%)  2 (2%)  1 (1%)  2 (2%) | 0.51 |
| Pre Injury Suicidality (%)  *No*  *Thoughts of self harm with no intent or plan*  *Thoughts of self harm with intent and/or plan*  *Single suicide attempt*  *Repeat suicide attempts* | 36 (82%)  4 (9%)  1(2%)  0 (0%)  3 (7%) | 102 (87%)  9 (8%)  5 (4%)  1 (1%)  0 (0%) | 0.06 |
| Median Glasgow Outcome Scale – Extended [IQR] | 8 [2.5] | 7 [2] | 0.77 |
| Abbreviations: COWAT Controlled Oral Word Association Test; CVLT California Verbal Learning Test; DTI diffusion tensor imaging | | | |

| **Supplementary Table 3**: Lesion presence and emotional wellbeing | | |
| --- | --- | --- |
| **Variable** | Adjusted $\beta$ | P value |
| Presence of Lesion | 4.59 [0.99, 8.18] | 0.014 |
| Age (per 10 years) | 0.93 [-0.52, 2.38] | 0.21 |
| Sex (ref: Female) | 0.45 [-3.56, 4.47] | 0.82 |
| Education (per 4 years) | -1.55 [-4.14, 1.04] | 0.24 |
| Time since injury (per 5 years) | 0.06 [-0.85, 0.98] | 0.89 |
| Pre-injury psychiatric history (yes/no) | -1.69 [-6.47, 3.10] | 0.49 |
| Overall cognitive composite score | 0.17 [-0.03, 0.38] | 0.10 |

| **Supplementary Table 4:** Lesion presence and Neuro-QOL Anxiety | | |
| --- | --- | --- |
| **Variable** | Adjusted $\beta$ | P value |
| Presence of Lesion | -3.76 [-6.41, -1.10] | 0.006 |
| Age (per 10 years) | -0.57 [-1.63, 0.50] | 0.30 |
| Sex (ref: Female) | -2.06 [-5.14, 1.02] | 0.19 |
| Education (per 4 years) | 0.04 [-1.78, 1.86] | 0.97 |
| Time since Injury (per 5 years) | 0.56 [-0.10, 1.23] | 0.10 |
| Pre-injury psychiatric history (yes/no) | -0.03 [-3.62, 3.55] | 0.98 |
| Abbreviations: QOL Quality of Life | | |

| **Supplementary Table 5:** Lesion presence and Neuro-QOL Depression | | |
| --- | --- | --- |
| **Variable** | Adjusted $\beta$ | P value |
| Presence of Lesion | -4.05 [-6.65, -1.45] | 0.003 |
| Age (per 10 years) | -0.71 [-1.74, 0.33] | 0.18 |
| Sex (ref: Female) | -0.28 [-3.31, 2.75] | 0.86 |
| Education (per 4 years) | -0.22 [-2.00, 1.55] | 0.81 |
| Time since Injury (per 5 years) | 0.36 [-0.28, 1.01] | 0.27 |
| Pre-injury psychiatric history (yes/no) | 1.10 [-2.55, 4.76] | 0.55 |
| Abbreviations: QOL Quality of Life | | |

| **Supplementary Table 6:** Lesion size and emotional wellbeing | | |
| --- | --- | --- |
| **Variable** | Adjusted $\beta$ | P value |
| Lesion Volume (per cc^3^) | 0.01 [-0.05, 0.08] | 0.72 |

| **Supplementary Table 7:** Global mean white-matter integrity and Neuro-QOL Anxiety | | |
| --- | --- | --- |
| **Variable** | Adjusted $\beta$ | P value |
| Mean white matter FA | 2.73 [1.02, 4.45] | 0.002 |
| Age (per 10 years) | 0.42 [-0.84, 1.67] | 0.52 |
| Sex (ref. Female) | -1.13 [-4.50, 2.24] | 0.51 |
| Education (per 4 years) | 0.68 [-1.31, 2.68] | 0.50 |
| Time since Injury (per 5 years) | 0.47 [-0.22, 1.16] | 0.19 |
| **Scanner** (ref: Ingenia)  *Skyra* | 6.15 [-1.93, 14.23] | 0.14 |
| CNR | -0.45 [-10.33, 9.42] | 0.93 |
| Absolute Motion (mm) | 3.73 [-0.55, 8.01] | 0.09 |
| Pre-injury psychiatric history (yes/no) | 1.18 [-3.01, 5.37] | 0.58 |
| Abbreviations: CNR Contrast to Noise Ratio; FA Fractional Anisotropy; QOL Quality of Life | | |

| **Supplementary Table 8:** Global mean white-matter integrity and Neuro-QOL Depression | | |
| --- | --- | --- |
| **Variable** | Adjusted $\beta$ | P value |
| Mean white matter FA | 2.29 [0.58, 4.01] | 0.010 |
| Age (per 10 years) | -0.08 [-1.35, 1.19] | 0.90 |
| Sex (ref. Female) | 1.12 [-2.28, 4.53] | 0.52 |
| Education (per 4 years) | 0.29 [-1.71, 2.29] | 0.78 |
| Time since Injury (per 5 years) | 0.35 [-0.34, 1.04] | 0.33 |
| **Scanner** (ref: Ingenia)  *Skyra* | 7.26 [-0.83, 15.35] | 0.08 |
| CNR | 0.05 [-9.86, 9.96] | 0.99 |
| Absolute Motion (mm) | 4.62 [0.32, 8.93] | 0.038 |
| Pre-injury psychiatric history (yes/no) | 3.53 [-0.88, 7.93] | 0.12 |
| Abbreviations: CNR Contrast to Noise Ratio; FA Fractional Anisotropy; QOL Quality of Life | | |

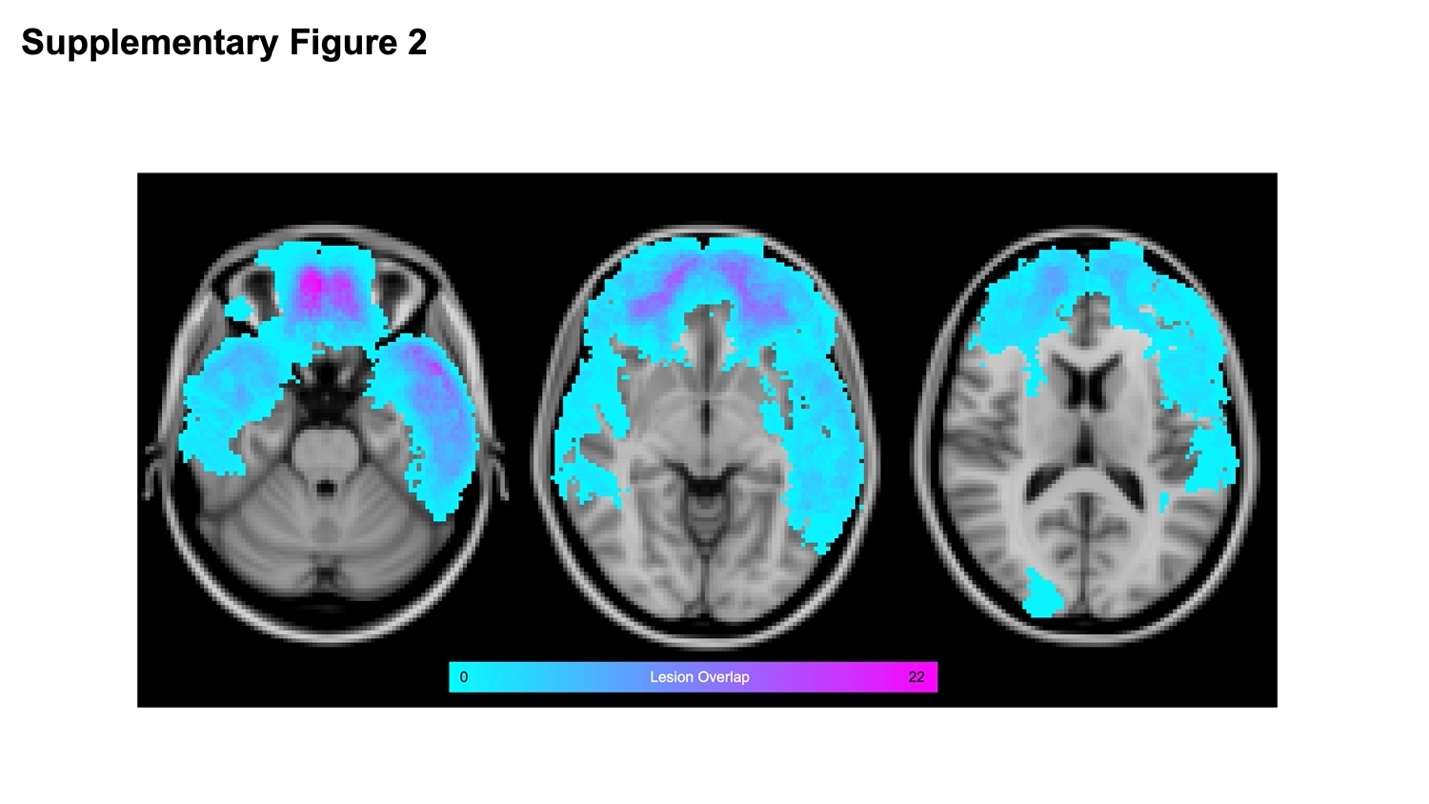

**Supplementary Figure 1: Lesion overlap map**

Distribution of encephalomalacic lesions in Montreal Neurological Institute T1 1mm Standard space. Color indicates absolute number of lesions at each voxel.

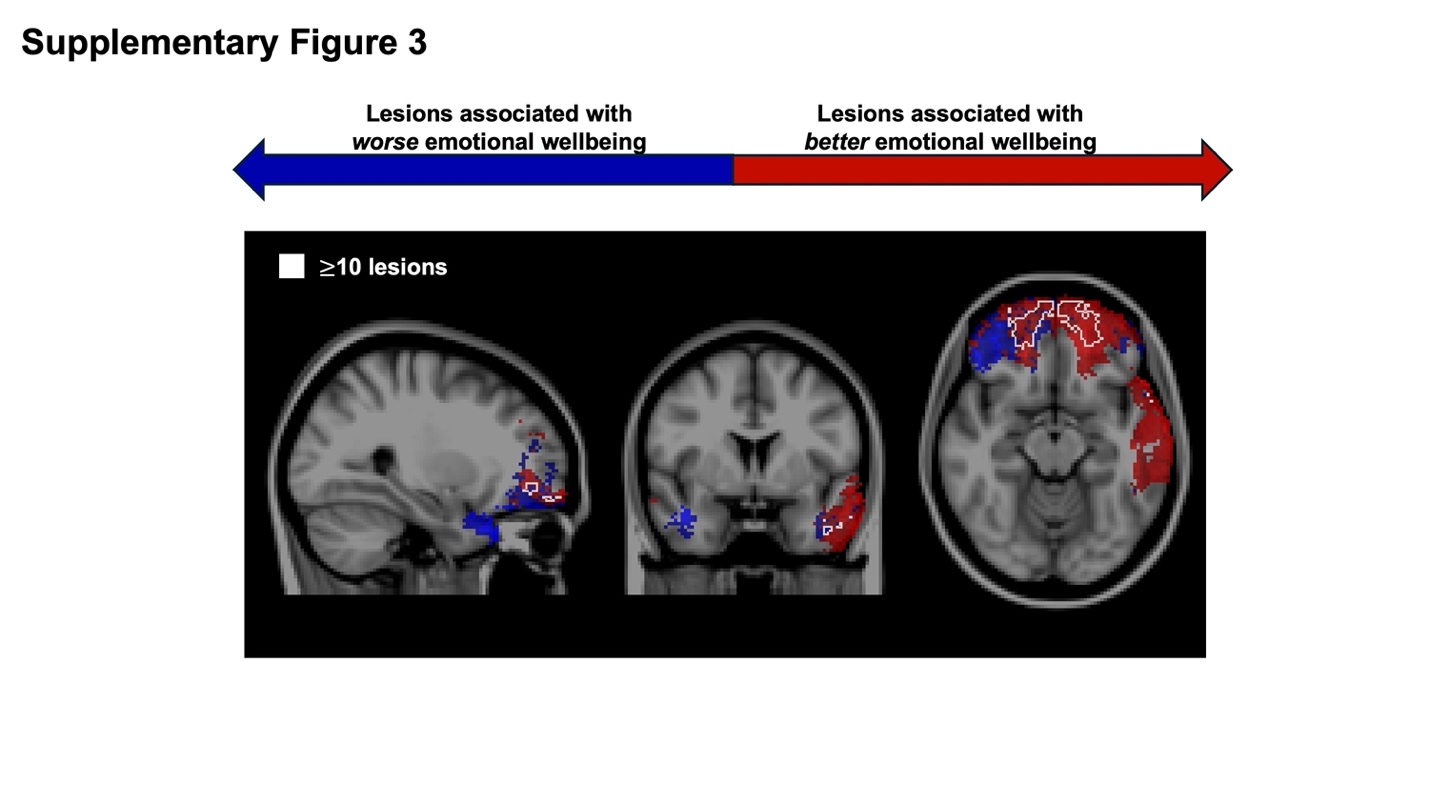

**Supplementary Figure 2: Lesion overlap map overlaid on voxel-wise associations with emotional wellbeing**

Voxels with at least 10 lesions are outlined in white. Voxels in red indicate voxels with a positive directional association emotional wellbeing (T>0) and voxels in blue indicate those with a negative association with emotional wellbeing (T<0).

**REFERENCES**

1.	Jonaitis EM, Koscik RL, Clark LR, et al. Measuring longitudinal cognition: Individual tests versus composites. Alzheimers Dement (Amst) 2019;11:74-84.

2.	Farrer TJ, Drozdick LW, John Wiley & Sons. Essentials of the California Verbal Learning Test : CVLT-C, CVLT-2, & CVLT3. In: Essentials of psychological assessment [online]2020.

3.	Wechsler D. Wechsler Adult Intelligence Scale—Fourth Edition (WAIS-IV). APA PsycTests 2008.

4.	Benton AL, Hamsher, d. S. K., & Sivan, A. B. Controlled Oral Word Association. APA PsycTests 1983.
